# Feature Integration of [^18^F]FDG PET Brain Imaging Using Deep Learning for Sensitive Cognitive Decline Detection

**DOI:** 10.64898/2026.01.22.26344669

**Authors:** Youjin Lee, Seonguk Kim, Sangil Kim, Yeona Kang, Alzheimer’s Disease Neuroimaging Initiative

## Abstract

**Background:** Distinguishing individuals with cognitive decline (CD), including early Alzheimer’s disease, from cognitively normal (CN) individuals is essential for improving diagnostic accuracy and enabling timely intervention. Positron emission tomography (PET) captures functional brain alterations associated with CD, but its broader application is often limited by cost and radiation exposure. To enhance the clinical utility of PET while addressing data limitations, we propose a multi-representational learning framework that leverages both imaging data and region-level quantification in a data-efficient manner.

**Methods:** Voxel-level features were extracted using convolutional neural networks (CNN) or principal component analysis networks (PCANet) from [¹⁸F]FDG PET imaging. Region-level features were derived from standardized uptake value ratio measurements across predefined brain regions and processed using a deep neural network (DNN). These voxel- and region-level information are integrated through direct concatenation. For final prediction, different machine learning models and ensemble technique were applied. The models were trained and validated using 5-fold cross-validation on PET scans from 252 participants in the Alzheimer’s Disease Neuroimaging Initiative (ADNI), comprising 118 CN and 134 CD subjects. Additional correlation analysis and disease classification comparison with the Mini-Mental State Examination (MMSE) were also performed.

**Results:** In 5-fold cross-validation, CNN, PCANet, and DNN models achieved classification accuracies of 0.69 ± 0.04, 0.69 ± 0.06, and 0.82 ± 0.06, respectively. The integrated DNN-CNN model using direct concatenation yielded the highest accuracy (0.87 ± 0.05), with a 6.10% improvement in accuracy and reduced standard deviation relative to the DNN-only model. Moreover, there were an increase of 14.29% in Recall (0.77 to 0.88) and an increase of 7.32% in F1-Score (0.82 to 0.88). Moreover, the model output showed a significant level of relation with MMSE, and it outperformed the MMSE-based classification in accuracy, recall, and f1, except precision.

**Conclusion:** Combining PET imaging with region-level quantification and deep learning improves diagnostic performance over single-feature based models. Notably, fusion-based approaches enhanced sensitivity to cognitive decline. This multimodal strategy offers a more data-efficient and accurate approach for classifying cognitive decline and supports broader PET application in clinical settings.

## Introduction

Alzheimer’s disease (AD) is a progressive neurodegenerative disorder that severely impairs cognitive function and activities of daily living (ADLs) [1–3]. As of March 2023, the World Health Organization estimates that over 55 million people worldwide are living with dementia, with AD being the most common form of dementia accounting for 60–70% of cases [4]. AD primarily affects brain regions responsible for memory, language, and reasoning, leading to a gradual decline independence and quality of life [1–3]. According to the Centers for Disease Control and Prevention, symptoms typically emerge after the age of 60, with the risk of AD doubles approximately every five years beyond age 65 [5, 6].

Mild cognitive impairment (MCI), often regarded as a precursor to AD [7, 8], involves noticeable cognitive decline that does not yet interfere significantly with ADLs [3]. However, over 50% of individuals with MCI progress to dementia within five years, with AD being the predominate cause [8, 9]. Evidences suggests that early intervention at the MCI stage may slow or even reverse the disease progression [3, 10]. Therefore, MCI related to AD represents a critical window for early diagnosis and treatment [11–16]. For this reason, AD and MCI are often grouped under the broader term “cognitively decline” (CD), highlighting the importance of early and reliable identification in both research and clinical setting.

Among current diagnostic tools, positron emission tomography (PET), particularly with [^18^F]fluorodeoxyglucose (FDG), plays a key role in the early detection of AD [17–19]. FDG PET enables the evaluation of cerebral glucose metabolism, revealing underlying neurodegenerative processes [20]. The standardized uptake value ratio (SUVr) remains the preferred quantification metric in both research and clinical practice due to its simplicity and robustness [21, 22].

Recent advances in deep learning have enhanced medical image analysis, including application in brain tumor segmentation using magnetic resonance imaging (MRI) [23], coronavirus disease 2019 detection from lung computed tomography scans [24], and AD classification (MRI and PET) [25]. Studies have explored diverse approaches to AD classification using multiple imaging modalities (e.g., MRI, fMRI, PET) and varying input levels (e.g., 3D ROI-based, 3D subject-level, 3D patch-level, and 2D slice-level data) [26, 27]. Hybrid models combining traditional machine learning (ML) with deep learning methods, as well as multi-institutional collaborations, have also been reported [27, 28].

For instance, a study using 2,552 PET scans from 836 participants conducted One-vs-All binary classification to distinguish each diagnostic group from the others, reporting classification accuracies of 0.74 (CN), 0.59 (MCI), and 0.78 (AD) [29]. Another study evaluated a model on 822 subjects (472 AD, 350 MCI) and achieved 0.79 accuracy on a hold-out test set (10% of the dataset) and 0.80 balanced accuracy during external validation [30]. Despite relatively limited datasets, these results highlight the growing feasibility of PET-based diagnostics, even in data-constrained clinical environments.

PET brain image offers a distinct advantage in detecting physiological changes earlier than structural imaging, offering valuable insights for diagnosis and disease monitoring [31, 32]. However, its clinical utility is often hindered by variations in scanner resolution and imaging protocols across institutions, which can introduce inconsistencies and limit the generalizability of analysis [33]. Models that perform well with small datasets could enhance PET’s clinical applicability across broader setting.

In our study, we present a deep learning framework to identify CD using a limited number of FDG PET scans. Our approach integrates both image-derived and region-based quantitative features from the same FDG PET modality, enabling more comprehensive analysis. We validated the performance of our feature fusion strategy across different dimensional levels and ML classifiers using data from the Alzheimer’s Disease Neuroimaging Initiative (ADNI). Finally, to provide insights into the interpretability of our models, we examined the relationship between the predicted probabilities and the clinical metrics, and compared their disease classification performance.

## Materials and Methods

### Data Collection and Characteristics

Data used in the preparation of this article were obtained from the Alzheimer’s Disease Neuroimaging Initiative (ADNI) database (adni.loni.usc.edu). The ADNI was launched in 2003 as a public-private partnership, led by Principal Investigator Michael W. Weiner, MD. The primary goal of ADNI has been to test whether serial MRI, PET, other biological markers, and clinical and neuropsychological assessment can be combined to measure the progression of MCI and early AD.

The research groups included subjects classified as CN, MCI, and AD. A total of 252 subjects were included in the study, consisting of 118 CN individuals and 134 patients, which included 70 individuals with MCI and 64 with AD. Moreover, one PET and one MRI scan were collected for each subject. Additional demographic and clinical variables, including sex, age, and the Mini-Mental State Examination (MMSE), were obtained when available.

The basic characteristic information of the dataset is stated in Table 1. Next, to avoid potential bias in the classification task, the distribution of key demographic variables, specifically sex and age, were examined across the dataset using statistical analysis. Moreover, to facilitate the clinical interpretation of our predictions, we analyzed whether there were significant differences in the distribution of MMSE between groups.

**Table 1.** Basic characteristic information of the dataset.

| Characteristic | Total | CN | CD |
| --- | --- | --- | --- |
| Total Subjects | 252<br>(100.0%) | 118<br>(46.8%) | 134<br>(53.2%) |
| Age, years | 75.5 ± 6.4 | 76.1 ± 5.6 | 75.1 ± 7.0 |
| Gender (M/F) | 132 / 120<br>(52.4% / 47.6%) | 61 / 57<br>(51.7% / 48.3%) | 71 / 63<br>(53.0% / 47.0%) |
| MMSE score | 26.3 ± 4.9<br>(N=147) | 29.2 ± 1.0<br>(N=76) | 23.2 ± 5.5<br>(N=71) |

The data are presented as mean ± SD or as n (%).

For PET imaging, we selected FDG PET scans. According to the PET protocols [34], the FDG tracer is administered at a dose of 185 MBq (5.0 mCi) with an allowable variation of ±10% across different studies (ADNI1, ADNI GO, ADNI2, ADNI3). The selected data are preprocessed including coregistered average, standard images, and voxel sizes with uniform resolution (described as “Coreg, Avg, Std Img and Vox Siz, Uniform Resolution”). During the preprocessing, cerebellar gray matter is used as the reference region for SUVr normalization. Thus, the collected PET imaging is an SUVr map, representing the relative FDG uptake compared to reference region.

For MRI imaging, T1-weighted images were collected within one month before or after the corresponding PET scan for the subject. This selection ensured that the MRI data was temporally aligned with the PET scans and MRI imaging is to conduct segmentation for region-based quantification analysis.

### Multi-representational Deep Learning Framework

The framework of our proposed multi-representational deep learning is shown in Fig 1. It consists of four phases: Data Preprocessing, Feature Extraction, Feature Fusion, and Classification. From a single imaging modality—FDG PET—two types of representations are derived during preprocessing: 2D PET slices and region-based quantification values. In the deep feature extraction stage, PET images and region-based quantification data are processed using separate deep learning architectures, each specialized for spatial or structured input respectively. The extracted features are integrated via direct concatenation at the feature fusion stage. The resulting fused feature vector is subsequently utilized as the input to the final ML-based classifier with ensemble technique for final prediction

**Figure 1.**
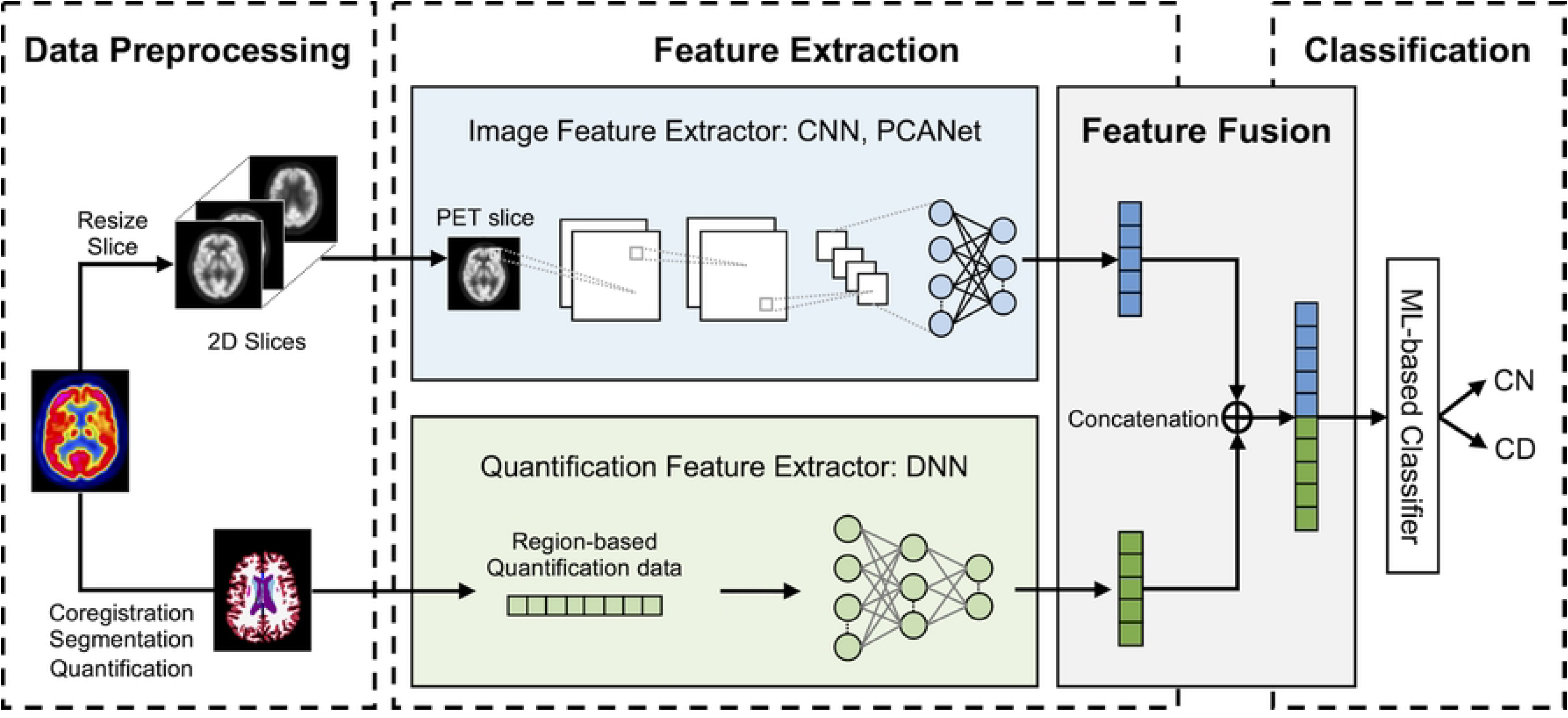
The framework of proposed multi-representational deep learning model.

### Data Preprocessing

The collected PET images were also utilized as inputs for deep learning models. Initially, the scans were resized to 128 × 128 × 64. To reduce the model size and the number of parameters required, the original 3D PET scans were sliced into 2D images. Specifically, 10 slices were extracted from the central region of the 3D PET scan, focusing on the ventricles, particularly in axial slices, for augmentation purpose during training (training and validation set). Additionally, only one middle slice was obtained for test set.

For region-based quantification analysis, segmentation, coregistration and quantification were conducted. Freesurfer [35] was used for the segmentation of each MRI image. The output of segmentation (aseg.mgz) divides the brain into 41 distinct regions, with detailed region label provided in Table S1. Each PET image was coregistered with the corresponding MRI image, and average activity for each region (SUVr value) extracted using the segmented mask. Both co-registration and quantification were processed on PMOD (version 3.8; PMOD Technologies Ltd [36]). Quantification data was collected for all subjects, resulting in tabular data representing region-based quantification.

To evaluate the model while ensuring its generalizability and robustness, we conducted 5-fold cross-validation. In each fold, the dataset was split into training, validation, and test sets in a ratio of 64%, 16%, and 20%, respectively. The split was conducted in subject level (the same as PET imaging level) and augmentation is applied after the split. Also, the same set division applied for PET imaging and quantification data.

### Feature Extraction

To integrate two different types of data, region-based quantification (tabular data) and 2D slice image (imaging data), deep learning models are employed to independently extract features from each data type. Deep Neural Networks (DNNs) were employed for tabular data, while Convolutional Neural Networks (CNNs) were utilized for imaging data due to their respective architectures and specialized capabilities, which align with the characteristics of these data types. To address the limitation of a small dataset, we utilized EfficientNetB0 [37], a compact and efficient CNN pretrained on ImageNet, and applied fine-tuning by unfreezing the last four layers to adapt the model to the target task. All detailed architecture of DNN and CNN is stated in Table S2 and Table S3.

Although the performance of CNNs is well recognized in vision tasks including various medical imaging tasks [38], it is also well known that a large size of dataset is required for high accuracy. As aiming to performance on smaller dataset, we employ simple neural network, Principal Component Analysis (PCA) Network (PCANet) [39]. PCANet follows similar stages as CNN; however, unlike CNNs, it does not require updating the weights of the filters. Instead, the filters are derived from PCA, eliminating the need for gradient-based optimization. This makes PCANet easier to implement and less computationally intensive, particularly for tasks with limited labeled data. All detailed setting for PCANet is stated in Table S4.

Specifically, the quantification data features were derived from three distinct DNN layers: DNN1 (with a feature dimensionality of 16), DNN2 (32), and DNN3 (64). Additionally, the original tabular data was incorporated into the fusion process and referred to as DNN0 (41). For PET imaging, both a convolutional neural network (CNN) and PCANet were employed. CNN features were extracted from three layers: CNN1 (with a dimensionality of 16), CNN2 (32), and CNN3 (1280). CNN3 corresponds to the final feature representation obtained from the pretrained EfficientNet-B0 model. In the case of PCANet, the extracted feature vector had a dimensionality of 128.

### Feature Fusion

The extracted features are subsequently fused in a concatenation manner. There are four feature dimension levels for each PET imaging and quantification data as described above and fusion is implemented in all possible 16 combination ways. Through concatenation, these features form a unified representation, enabling the model to leverage the complementary information contained within both PET imaging and quantification data.

### Classification

ML based classifiers were used to make the final prediction. A total of 12 machine learning classifiers were employed in this study, including Random Forest (RF), Extra Trees (ET), Multi-Layer Perceptron (MLP), Naive Bayes (NB), Gradient Boosting (GB), Logistic Regression (LR), k-Nearest Neighbors (KNN), Support Vector Machine (SVM), and Decision Tree (DT), all implemented using the Scikit-learn library [40]. In addition, eXtreme Gradient Boosting (XGBoost) [41], Light Gradient Boosting Machine (LightGBM) [42], and Categorical Boosting (CatBoost) [43] were utilized via their respective official Python libraries. To further boost model performance, to enhance robustness, we implemented ensemble methods by combining heterogeneous classifiers from different algorithmic families, including RF (tree-based), MLP (neural network), CB (gradient boosting), SVM (kernel-based), and LR (linear model). Ensemble strategies such as hard voting, soft voting, and stacking were applied using the predicted probabilities from each model.

### Model Assessment

Our proposed frameworks and baseline models were compared using evaluation metrics such as precision, recall, F1 score, accuracy, and the area under the ROC curve (AUC). All performance metrics were evaluated using 5-fold cross-validation and reported as the mean and standard deviation across folds. For confusion matrix analysis, predictions from all five folds were aggregated to allow for a more stable comparison across models. Among various evaluation metrics, we selected the F1-score as the primary performance indicator, as it provides a balanced assessment of precision and recall as shown in Eq (1), which serves as a more informative metric in clinical classification tasks due to its sensitivity to both false positives (FP) and false negatives (FN) beyond simply considering true positives (TP) and true negatives (TN) [44].

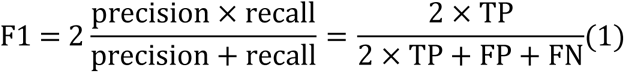

### Interpretability Analysis

Beyond binary classification, we clinically evaluate our predictions. Each model generates an output as a probability between 0 and 1, which is converted to 0 or 1 using a threshold of 0.5 for final prediction. We assume that the predicted probability may reflect the severity of CD. To examine this, we calculate the Pearson correlation coefficient (r) with MMSE, which is one of well-known cognitive assessment tool for evaluating various cognitive disorders [45].

Furthermore, we compared the discriminative ability between the models and MMSE. To accomplish this, we re-evaluated model performance using only the patients for whom clinical data were available (n = 147). Here, to classify individuals using MMSE, we categorized those with scores less than 24 as CD and those with scores greater than or equal to 24 as CN. For this analysis, we used the same predictions generated during the original 5-fold cross-validation and computed the performance metrics using all available predictions.

## Results

### Data Characteristics Analysis

The distribution of age for both CN and CD can be considered normally distributed since p-value from the Shapiro-Wilk normality test is greater than 0.05. With the assumption of normality, we conducted t-test and it showed no significant difference in the age distribution between CN and CD (p-value = 0.20). By chi-square test for sex distribution, we observed that there is no significant difference in the distribution of sex between CN and CD (p-value = 0.94). Thus, our dataset constructed for classification task dose not exhibit any bias with respect to sex or age.

Total 147 subjects out of 252 subjects are available for MMSE analysis. Firstly, we observed non-normality in the distribution of MMSE in both groups from the Shapiro-Wilk normality test (p < 0.001). So, the Mann-Whitney U test was employed as non-parametric alternative. According to the Mann-Whitney U test with a one-sided alternative hypothesis (greater), CN had significantly higher MMSE score than CD (p < 0.001). As a well-established clinical indicator, MMSE showed a statistically significant difference and is thus considered a representative clinical metric for further analysis.

### Baseline Models Performance

Firstly, the performance of baseline model is examined. DNN is employed for quantification data (tabular data) while CNN and PCANet are utilized for PET imaging. Since PCANet is only a feature extractor, 12 ML classifiers are employed to make the final prediction (Table S5). Among them, XGB shows the highest average F1-score - the primary evaluation metric - and showed superior results across all other metrics except for recall. Table 2 presents the performance of the three baseline models. Among baseline models, DNN achieved the best performance across all metrics, with an average F1-score of 0.82. and PCANet with XGB exhibit similar accuracy. However, CNN, like DNN, achieves higher precision, while PCANet shows a substantially higher recall.

**Table 2.**
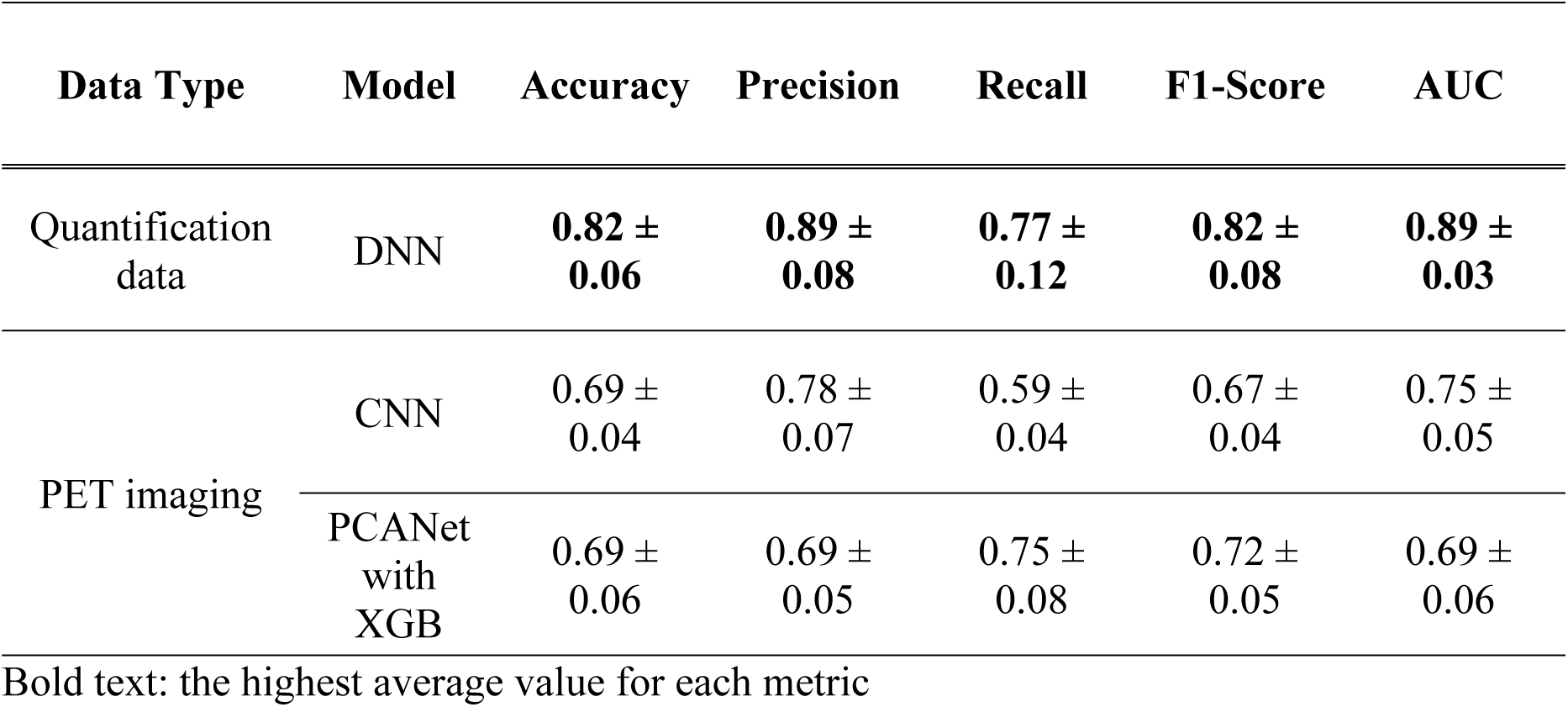
Comparison of Baseline model – DNN, CNN, PCANet with XGB.

### Fusion Model Performance Comparison

To investigate the performance models across various combinations of feature levels and final classifiers, we firstly fixed classifier to RF, which shows the most frequently best-performing classifier, and varied the feature levels. Overall, DNN2 has higher average accuracy of 0.86 with standard deviation 0.01 (DNN0: 0.81 ± 0.02, DNN1: 0.85 ± 0.01, DNN3: 0.084 ± 0.01) for quantification data. In the case of PET imaging, CNN3 has slightly higher average accuracy of 0.85 with standard deviation 0.02 (PCANet: 0.84 ± 0.01, CNN1: 0.84 ± 0.02, CNN3: 0.83 ± 0.03) as shown in Table 3. The highest F1-score shows 0.87 with the combination of (DNN2, CNN1) and (DNN2, CNN2) (Table 3). They also outperformed in other metrics excepts, and the evaluation results with all metrics for all combinations are reported in Table S6.

**Table 3.** Average F1-score of each feature levels with RF.

|  | **DNN0** | **DNN1** | **DNN2** | **DNN3** | **AVG** | **SD** |
| --- | --- | --- | --- | --- | --- | --- |
| **PCANet** | 0.84 | 0.84 | 0.85 | 0.84 | 0.84 | 0.01 |
| **CNN1** | 0.81 | 0.85 | **0.87** | 0.84 | 0.84 | 0.02 |
| **CNN2** | 0.82 | 0.86 | **0.87** | 0.85 | **0.85** | 0.02 |
| **CNN3** | 0.78 | 0.85 | 0.86 | 0.82 | 0.83 | 0.03 |
| **AVG** | 0.81 | 0.85 | **0.86** | 0.84 |  |  |
| **SD** | 0.02 | 0.01 | 0.01 | 0.01 |  |  |

(DNN# and CNN# show different layer types)

Next, fixing the latent variable dimension for (DNN2, CNN1), we conducted a comparative analysis of all ten ML classifiers. Also, ensemble approaches using 5 ML models (RF, MLP, CB, SVM, and LR), denoted by Ensemble, were applied with soft voting, stacking, and hard voting. As shown in Table S7, RF outperforms other individual classifiers in all evaluation metrics including a F1-score of 0.87 among single ML classifiers. While the ensemble approach yields marginally increasing in all metrics: Accuracy (0.86 to 0.87), Precision (0.88 to 0.89), Recall (0.86 to 0.88), F1-scores (0.87 to 0.88) and AUC (0.86 to 0.88) (Table S8).

### Class-wise Performance Analysis

All baseline and fusion, ensemble models are summarized in Table 4. We denoted the fusion model using RF with features from DNN2 and CNN1 as Fusion 1 and the combination of DNN2 and CNN2 as Fusion 2. Corresponding best ensemble models are referred to as Ensemble 2 and Ensemble 3.

**Table 4.**
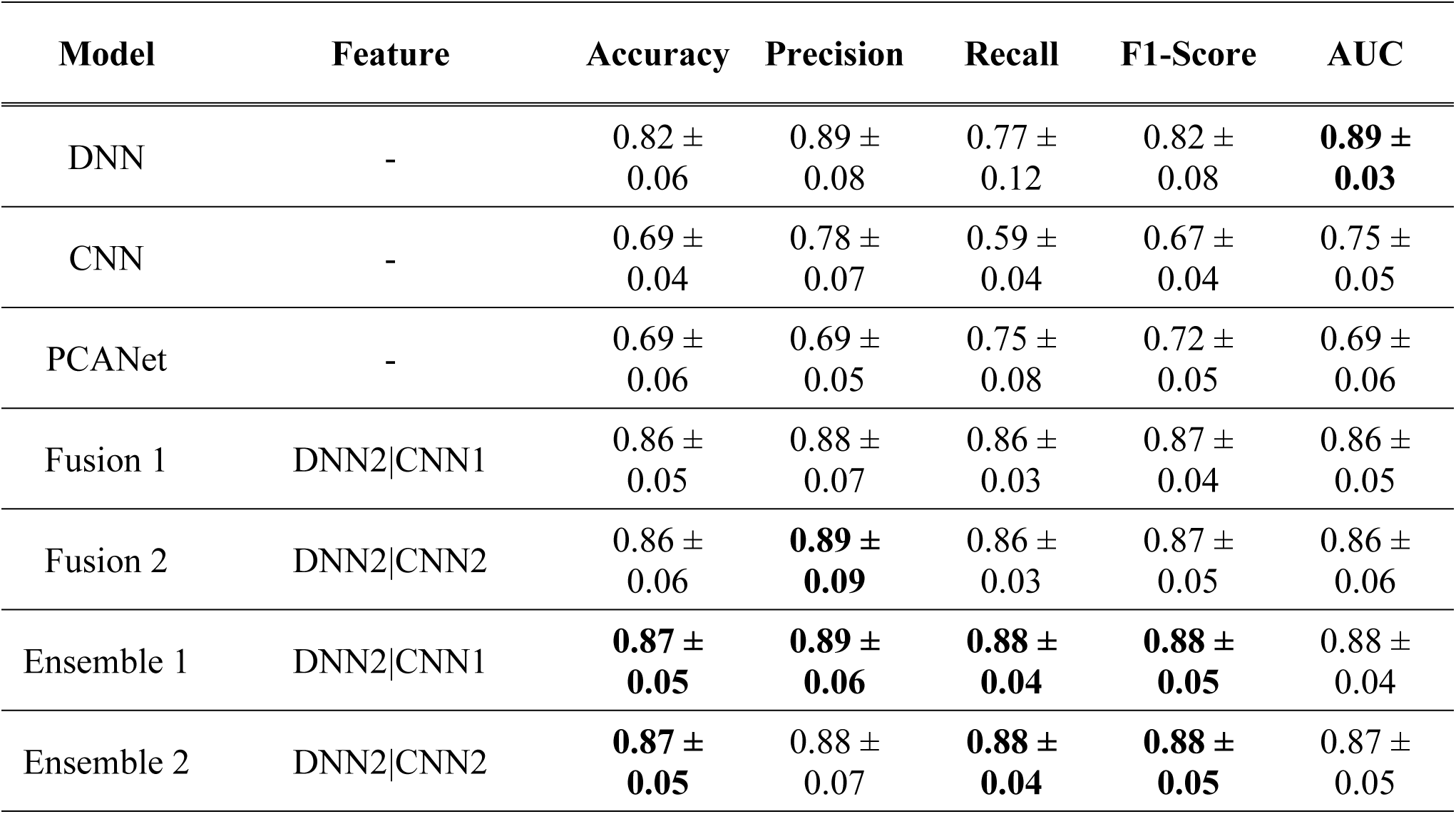
Summary of baseline, fusion, and ensemble models’ performance.

| Model | Feature | Accuracy | Precision | Recall | F1-Score | AUC |
| --- | --- | --- | --- | --- | --- | --- |
| DNN | - | 0.82 ± 0.06 | 0.89 ± 0.08 | 0.77 ± 0.12 | 0.82 ± 0.08 | **0.89 ± 0.03** |
| CNN | - | 0.69 ± 0.04 | 0.78 ± 0.07 | 0.59 ± 0.04 | 0.67 ± 0.04 | 0.75 ± 0.05 |
| PCANet | - | 0.69 ± 0.06 | 0.69 ± 0.05 | 0.75 ± 0.08 | 0.72 ± 0.05 | 0.69 ± 0.06 |
| Fusion 1 | DNN2\|CNN1 | 0.86 ± 0.05 | 0.88 ± 0.07 | 0.86 ± 0.03 | 0.87 ± 0.04 | 0.86 ± 0.05 |
| Fusion 2 | DNN2\|CNN2 | 0.86 ± 0.06 | **0.89 ± 0.09** | 0.86 ± 0.03 | 0.87 ± 0.05 | 0.86 ± 0.06 |
| Ensemble 1 | DNN2\|CNN1 | **0.87 ± 0.05** | **0.89 ± 0.06** | **0.88 ± 0.04** | **0.88 ± 0.05** | 0.88 ± 0.04 |
| Ensemble 2 | DNN2\|CNN2 | **0.87 ± 0.05** | 0.88 ± 0.07 | **0.88 ± 0.04** | **0.88 ± 0.05** | 0.87 ± 0.05 |

Compared to DNN – the best performing baseline model, both fusion and ensemble models show improved performance. Overall, the average performance become better with smaller standard deviation as a result of feature integration. With precision maintained at a similar level, the substantial improvement in recall indicates that the model has become more sensitive to true positive (CD) detection.

According to the aggregate confusion matrix in Fig. 2, both DNN and CNN models demonstrate strong performance in predicting true negatives (CN), with accuracy of 0.88 and 0.81, respectively, whereas PCANet shows a lower prediction in this regard. Although PCANet shows the higher accuracy in true positives (CD) compared to its performance on true negatives.

**Figure 2.**
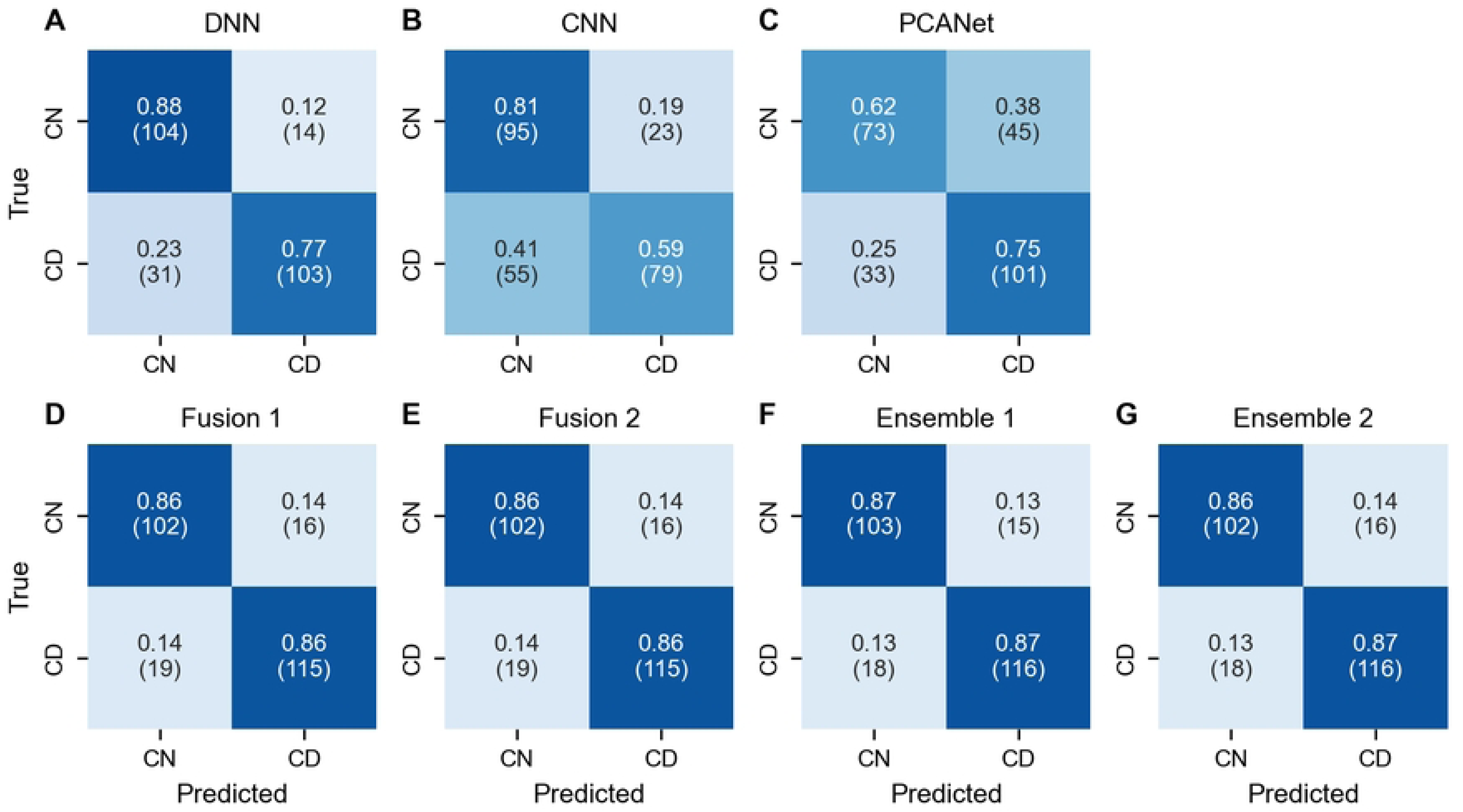
Confusion matrix for baseline, fusion, and ensemble models.

Fusion and Ensemble models achieved both true negatives and true positives above 0.85, showing an increase of at least 0.08 in true positive compared to DNN. Among these models, Ensemble 1, fusing DNN2 and CNN1 with ensemble ML classifier, can be considered as the optimal model with an average F1-score 0.88, true negatives 0.88, and true positive 0.88.

### Interpretability Analysis

Moreover, the correlation analysis between MMSE and predicted probabilities from each model was conducted on test set (Figure 3). Total 147 subjects of out 252 subjects in all test set during 5-fold cross validation were included. From the analysis, we observed moderate negative relations (−0.8 < r < −0.5) except the probabilities from PCANet, having weak relation. However, since all correlations have p < 0.001, there is a significant linear relationship between MMSE scores and predicted probabilities.

**Figure 3.**
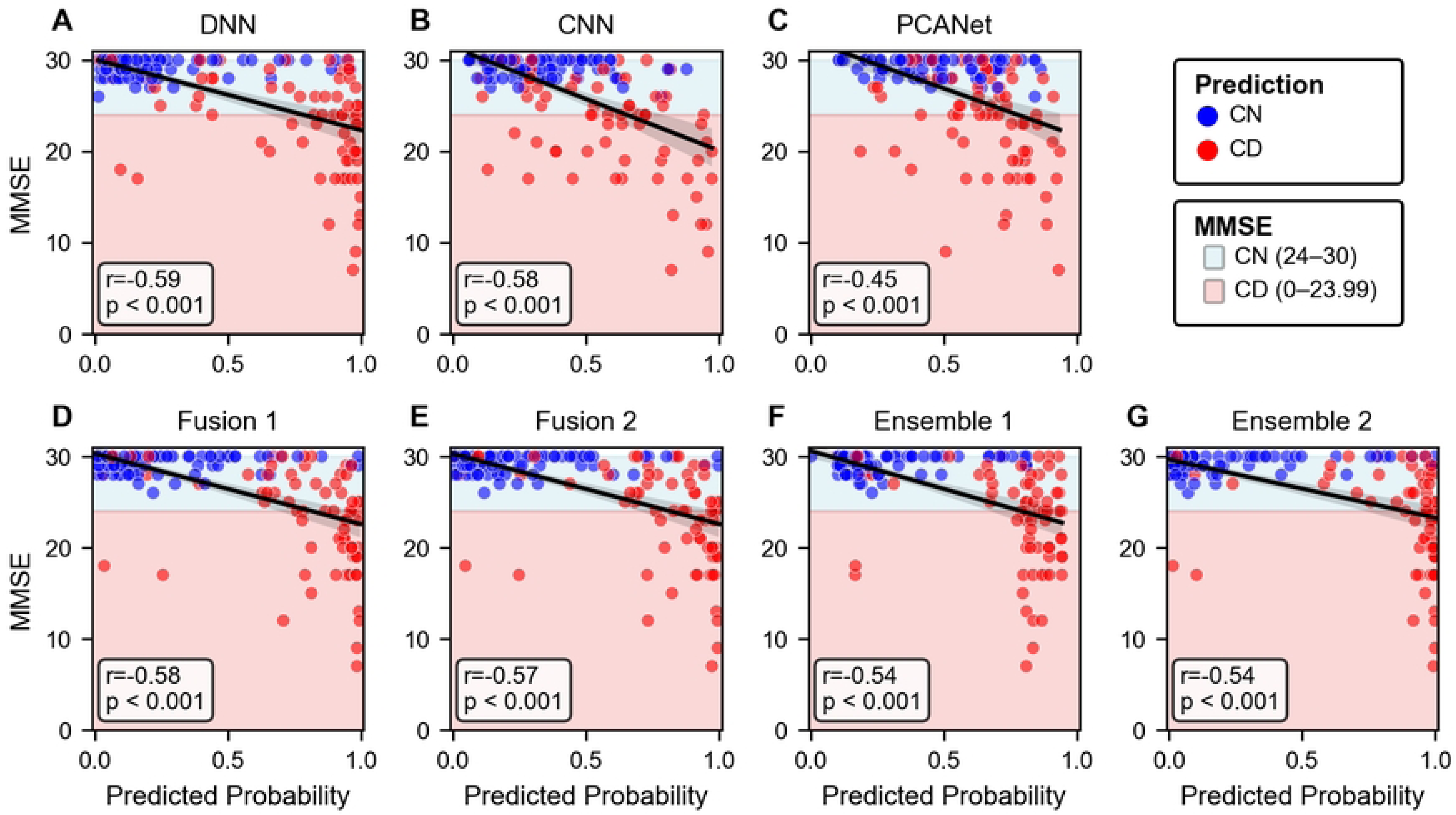
Correlation between MMSE and predicted probabilities from each model.

In the classification comparison between MMSE and our optimal model (Ensemble 1), Ensemble model shows higher performance except precision across the metrics (Table 5). MMSE has strong ability in correctly identifying CN (higher precision), while our model demonstrates superior performance in correctly detecting CD (higher recall). Similar results were obtained when performance was summarized as the average across folds (Table S9).

**Table 5.** Comparison of classification by optimal model and MMSE.

| Model | Accuracy | Precision | Recall | F1-Score | AUC |
| --- | --- | --- | --- | --- | --- |
| Ensemble 1 | 0.88 | 0.86 | 0.89 | 0.88 | 0.91 |
| MMSE | 0.72 | 1.00 | 0.42 | 0.59 |  |

## Discussion

Although many recent studies on Alzheimer’s disease diagnosis have focused on MRI due to its higher spatial resolution, PET provides complementary molecular-level information that reflects underlying disease processes such as glucose metabolism. This makes PET particularly valuable for identifying cognitive decline, even when structural changes are less pronounced. In our study, we developed a deep learning framework that leverages FDG-PET by integrating both image-derived and region-based quantitative features, enabling a more comprehensive representation of disease-related patterns. By validating this feature fusion approach across different dimensional levels and classifiers, we demonstrate that multi-feature modeling approaches improve the classification performance, especially in CD sensitivity, which regards more critical in clinical setting.

Fusion model with ensemble methods achieved the highest performance with accuracy of 0.87, recall of 0.88, and F1-score of 0.88. However, the tabular data based DNN still shows good performance with accuracy of 0.82 and it has the highest score in precision and AUC, which implies the region-based quantification has solid physiological information and also indicates the high contribution overall integration models. Nevertheless, the integration approaches still show the potential of improving in CD sensitivity (recall: 0.77 to 0.88) and more robust performance with lower standard deviation in 5-fold cross validation.

In the analysis with MMSE, there was a moderate relation between models and MMSE, but the absolute correlation coefficient was not correlated with the model performance. Ensemble 1, which showed the best performance, did not have the highest correlation. This is because the CD has a wide range of MMSE, so even good CD detection could not have a high correlation with MMSE. Therefore, we examined the performance comparison with the diagnosis using MMSE. In the case of MMSE, the recall was relatively low while the precision was 1.00. This implies that MMSE does not capture the CD at the early stage. In clinical setting, the final diagnosis is made using multiple sources of information, not solely based on MMSE. However, the output of our model may have potential as a diagnostic aid, especially given its sensitivity to CD.

In our study, there are several limitations. First, many models are involved in our frameworks, so it is complicated to make the optimal choice for hyperparameters related to models and we compared the models in general setting. In other words, the performance could potentially be further improved through hyperparameter tuning, such as grid search or simulated annealing. Next, since our study required the segmentation of each scan and region-based quantification, relatively small sized dataset was available, which better reflects real-world clinical settings. To ensure a transparent and reliable performance evaluation on a small dataset, we conducted 5-fold cross-validation and additionally reported both the mean and standard deviation.

Although there are many advantages of PET, PET has lower resolution, and it is a challenging for PET-based model to achieving the higher accuracy comparing other modality (such as MRI) derived models. To tackle this challenging, several postprocessing strategies could be explored in future work to enhance resolution.

Additionally, alternative feature concatenation methods, such as canonical correlation analysis (CCA) could be explored to further improve performance. Although CCA was tested in our experiments, it did not yield meaningful improvement, possibly due to suboptimal dimensional configurations. A more systematic tuning across dimensions may reveal its potential in future work.

## Conclusions

This study presented a deep learning framework that integrates image-derived and region-based features from FDG PET data to classify cognitive decline. Fusion and ensemble models achieved superior performance, particularly improving recall, which is critical for early detection of Alzheimer’s disease. Correlation with MMSE scores further confirmed the clinical relevance of our predictions and beyond this our model shows the better disease differentiability compared to MMSE. These findings suggest that integrating distinct PET-derived representations improves diagnostic accuracy and interpretability, indicating a promising direction for machine learning–based approaches in Alzheimer’s disease diagnosis and monitoring.

## Author Contributions

Conceptualization: Youjin Lee, Yeona Kang

Data curation: Youjin Lee, Seonguk Kim

Formal analysis: Youjin Lee

Funding Acquisition: Sangil Kim, Yeona Kang

Investigation: Youjin Lee

Methodology: Youjin Lee, Seonguk Kim

Project administration: Yeona Kang

Resources: Sangil Kim, Yeona Kang

Software: Youjin Lee, Seonguk Kim, Yeona Kang

Supervision: Yeona Kang

Validation: Youjin Lee

Visualization: Youjin Lee

Writing – original draft: Youjin Lee

Writing – review & editing: Youjin Lee, Seonguk Kim, Yeona Kang

## Acknowledgements

Data collection and sharing for this project was funded by the Alzheimer’s Disease Neuroimaging Initiative (ADNI) (National Institutes of Health Grant U01 AG024904) and DOD ADNI (Department of Defense award number W81XWH-12-2-0012). ADNI is funded by the National Institute on Aging, the National Institute of Biomedical Imaging and Bioengineering, and through generous contributions from the following: AbbVie, Alzheimer’s Association; Alzheimer’s Drug Discovery Foundation; Araclon Biotech; BioClinica, Inc.; Biogen; Bristol-Myers Squibb Company; CereSpir, Inc.; Cogstate; Eisai Inc.; Elan Pharmaceuticals, Inc.; Eli Lilly and Company; EuroImmun; F. Hoffmann-La Roche Ltd and its affiliated company Genentech, Inc.; Fujirebio; GE Healthcare; IXICO Ltd.; Janssen Alzheimer Immunotherapy Research & Development, LLC.; Johnson & Johnson Pharmaceutical Research & Development LLC.; Lumosity; Lundbeck; Merck & Co., Inc.; Meso Scale Diagnostics, LLC.; NeuroRx Research; Neurotrack Technologies; Novartis Pharmaceuticals Corporation; Pfizer Inc.; Piramal Imaging; Servier; Takeda Pharmaceutical Company; and Transition Therapeutics. The Canadian Institutes of Health Research is providing funds to support ADNI clinical sites in Canada. Private sector contributions are facilitated by the Foundation for the National Institutes of Health (www.fnih.org). The grantee organization is the Northern California Institute for Research and Education, and the study is coordinated by the Alzheimer’s Therapeutic Research Institute at the University of Southern California. ADNI data are disseminated by the Laboratory for Neuro Imaging at the University of Southern California.

## Supporting information

## Data Availability

The data underlying the results presented in this study were obtained from the Alzheimer’s Disease Neuroimaging Initiative (ADNI) database (https://adni.loni.usc.edu/).

